# Predicting mortality of individual COVID-19 patients: A multicenter Dutch cohort

**DOI:** 10.1101/2020.10.10.20210591

**Authors:** Maarten C. Ottenhoff, Lucas L. Ramos, Wouter Potters, Marcus L.F. Janssen, Deborah Hubers, Dan Piña-Fuentes, Rajat Thomas, Iwan C.C. van der Horst, Christian Herff, Pieter Kubben, Paul W.G. Elbers, Henk A. Marquering, Max Welling, Shi Hu, Suat Simsek, Martijn D. de Kruif, Tom Dormans, Lucas M. Fleuren, Michiel Schinkel, Peter G. Noordzij, Joop P. van den Bergh, Caroline E. Wyers, David T. B. Buis, Joost Wiersinga, Ella H.C. van den Hout, Auke C. Reidinga, Daisy Rusch, Kim C.E. Sigaloff, Renée Douma, Lianne de Haan, Egill A. Fridgeirsson, Niels C. Gritters van den Oever, Roger J.M.W. Rennenberg, Guido A. van Wingen, Marcel J.H. Aries, Martijn Beudel, on behalf of The Dutch COVID-PREDICT research group

## Abstract

**Objective:** Develop and validate models that predict mortality of SARS-CoV-2 infected patients admitted to the hospital.

**Design:** Retrospective cohort study

**Setting:** A multicenter cohort across ten Dutch hospitals including patients from February 27 to June 8 2020.

**Participants:** SARS-CoV-2 positive patients (age ≥ 18) admitted to the hospital.

**Main Outcome Measures:** 21-day mortality evaluated by the area under the receiver operatory curve (AUC), sensitivity, specificity, positive predictive value and negative predictive value. The predictive value of age was explored by comparison with age-based rules used in practice and by excluding age from analysis.

**Results:** 2273 patients were included, of whom 516 had died or discharged to palliative care within 21 days after admission. Five feature sets, including premorbid, clinical presentation and laboratory & radiology values, were derived from 80 features. Additionally, an ANOVA-based data-driven feature selection selected the ten features with the highest F-values: age, number of home medications, urea nitrogen, lactate dehydrogenase, albumin, oxygen saturation (%), oxygen saturation is measured on room air, oxygen saturation is measured on oxygen therapy, blood gas pH and history of chronic cardiac disease. A linear logistic regression (LR) and non-linear tree-based gradient boosting (XGB) algorithm fitted the data with an AUC of 0.81 (95% confidence interval 0.77 to 0.85) and 0.82 (0.79 to 0.85), respectively, using the ten selected features. Both models outperformed age-based decision rules used in practice (AUC of 0.69, 0.65 to 0.74 for age > 70). Furthermore, performance remained stable when excluding age as predictor (AUC of 0.78, 0.75 to 0.81)

**Conclusion:** Both models showed excellent performance and had better test characteristics than age-based decision rules, using ten admission features readily available in Dutch hospitals. The models hold promise to aid decision making during a hospital bed shortage.

## INTRODUCTION

The first wave of the COVID-19 pandemic had a dramatic effect on our society and severely disrupted our daily lives, economies and healthcare systems. During the peak of the first wave, hospitals and intensive care units (ICU) throughout Europe were overwhelmed and resources were exhausted. Implementation of public health policies reduced the infection-rate; however, there is a considerable risk that relaxation of these policies lead to a second pandemic wave, of which the first signs are already seen in different European countries.[1] The progression of the Spanish flu pandemic learned that a second wave could impose an even higher demand on the healthcare system and result in a higher number of casualties than the first wave.

Given the novelty of the virus, accurate information about the clinical course and prognosis of individual patients is still largely unknown, which led to the use of crude limits to unilaterally withhold advanced life support measures to face the large numbers of pulmonary insufficient patients during the first wave. Although criticized, several hospitals in Europe have already solely used age as a triage criterion.[2] Many publications have developed and evaluated triage selection criteria, but a there remains a significant knowledge gap and the final criteria are subject to socio-ethical debate.[3–5] Preferably, triage is averted, but when necessary, the decision should be based on medical criteria with an evidence base. Since March 2020, many studies have been published regarding the clinical characteristics of patients suffering from a SARS-CoV-2 infection in both smaller (n=58 [6], n=200 [7]) and larger cohorts (n > 5000 [8– 10]). However, these studies have reported notable differences in clinical characteristics that were associated with an adverse outcome. Importantly, these studies only provide information about clinical characteristics and risk factors on group level, and therefore do not provide information about the prognosis for individual patients. To prevent reuse of crude limits when hospital and ICU resources are exhausted during a second wave, a prognostic model using multivariable analysis could be of great value. Such a model can provide information about the individual patients’ chance of survival, despite largely unknown underlying risk factors.

Within the ongoing socio-ethical debate in the Netherlands, whether age should be included in the triage selection criteria, such a predictive model could allow to exclude age or include it in a combination with clinical characteristics. All of the published prognostic are continuously reviewed by Wynants et al. 2020.[11], who identified 145 prediction models of which 23 were tailored towards predicting mortality. The authors identified that all studies were at high risk of bias and likely to underperform in clinical practice. However, a recent paper, not yet reviewed by Wynants et al., showed promising results on predicting mortality with excellent performance, using a very large cohort (n > 50.000) from the United Kingdom.[12]

The uncertainty and risk of bias in almost all published COVID-19 related prognostic models, stresses the importance of thorough methodology in variable selection, internal and external model validation and performance evaluation.[11] In addition, it is important that a constant interplay between data-scientists and clinicians is in place during model development.

Furthermore, studies developed and performed independently with similar methodology are more valuable than ever to reduce the uncertainty of published models and risk of spurious publications.[13,14] Therefore, a prognostic model was developed and evaluated that predicts 21-day mortality; utilizing data from 2273 SARS-CoV-2 infected patients from 10 hospitals across the Netherlands.

## MATERIALS AND METHODS

### Data collection

Data were included from 10 Dutch hospitals varying from small to large peripheral hospitals to large academic centers. For an up-to-date overview of the including centers, see www.covidpredict.org. Clinical data were derived from electronic health records, pseudonymised and stored in an online database (Castor EDC, Amsterdam, The Netherlands) by each hospital independently. Data collection started with the first admitted patients in the including centers. This was after the first confirmed case in the Netherlands on February 27, 2020. Records were included to an admission date up until June 8th when the Dutch admission rates sharply decreased.[15] Inclusion criteria were admission in a hospital, age >= 18 years, a positive SARS-CoV-2 PCR before or during admission, or a CO-RADS CT thorax score >= 4 at admission. Retrospective data collection was based on the rapid COVID-19 case report form (rCCRF) developed by the WHO.[16] After consultation with several specialist consultants and an evaluation of the COVID-19 literature (mainly from China and Italy), additional clinical and laboratory features were added to the rCCRF.

The study protocol was reviewed by the medical ethics committees of the Amsterdam University Medical Centers (Amsterdam UMC; 20.131) and Maastricht University Medical Center (MUMC; 2020-1323). Given the exceptional circumstances related to the COVID-19 crisis and in accordance with national guidelines and European privacy law, the need for informed consent was waived and opt out procedure was communicated by press release.

Despite this, individual centers used local guidelines to obtain consent retrospectively from patients or representatives. In all centers, measures were taken to ensure adequate and safe data pseudonymisation and storage.

### Outcome definition

To support the decision of (ICU) treatment during scarcity at hospital admission, we aim to predict unfavorable outcome of COVID-19 patients at hospital admission. Given the amount of data, predicting each possible outcome, such as mortality, palliative care, discharge, and hospitalized, could increase the risk of biased models and overfitting. Therefore, the prediction goal was modelled as a binary classification problem, where an unfavorable outcome corresponds to patients that either died or were discharged for palliative care within 21 days after hospital admission. Palliative discharge is end-of-life care that focuses on patient comfort rather than treatments with curative intentions. A favorable outcome corresponds to patients that are discharged to home, nursing homes or rehabilitation units within 21 days and patients that are alive and still hospitalized at 21 days after hospital admission. Patients that were still hospitalized but shorter than 21 days, transferred to other hospitals (including transfers to participating hospitals), re-admitted or have an unknown outcome were excluded from further analysis.

### Data processing and quality

The rCCRF was filled in manually by a large team of researchers and doctors the electronic patient dossiers in the different hospitals could not be coupled to the Castor database. The rCCRF and additional features resulted in a large amount of features (> 400). A consensus meeting with clinicians was held (April 18^th^, 2020) to remove features that were not available at hospital admission, not within the standard admission laboratory values or at risk of bias. This resulted in a feature set of 80 features. These 80 features were then divided into 6 sets: (1) premorbid characteristics (n=24), (2) clinical characteristics at admission (n=14), (3) laboratory and radiology findings at admission (n=42), (4) the combination of set 1 and 2 (n=38), (5) all features (n=80) and (6) a data-driven selection from all features (n=10). The process of the data-driven selection is described further in the modelling process section. A complete overview of all features per set is shown in supplementary table 1 and numerical characteristics per set is shown in supplementary table 4. The resulting features were checked for physiologically implausible outliers by two authors (MA/DH). Some features contained high but plausible values and were therefore not removed (e.g. creatinine kinase). Furthermore, collinearity was assessed by a Pearson correlation matrix.

### Predictive modelling

The obtained models could ultimately change the clinicians’ decision and thus directly influences the life of a patient. It is therefore of utmost importance that the obtained models are both robust and interpretable.[17] To comply with these requirements, two models with a fundamentally different modelling approach were selected: a logistic regression (LR) that fits the data linearly, and a tree-based gradient boosting algorithm that fits the data non-linearly. The models were implemented using the python 3 libraries Scikit-learn[18] and XGBoost (XGB)[19], respectively. Both models can be interpreted relatively easy and XGB often shows state-of-the-art results in multiple tasks. The models were trained and validated using leave-one-hospital-out cross-validation (LOHO-cv). By iteratively training the models on all but one hospital and testing performance on the left-out hospital, the performance of the model represents the ability to predict the outcome on independent data and thereby incorporate possible data heterogeneity between hospitals. To prevent skewed performance on individual folds due to a small number of samples, we combined the data from the two hospitals with the smallest number of samples and considered them as a single hospital in LOHO-cv for further analysis. Additionally to LOHO-cv, internal 10-fold random subsampling cross-validation using data from all hospitals was performed to facilitate a comparison of the results to other studies that typically only perform internal cross-validation.

### Modelling process

Features that had more than 50% missing values and subsequently patient records that had more than 80% missing values were removed. The remaining missing values were imputed using a multivariate iterative imputer (trained on the training set and applied to the test set). The imputer, a Bayesian ridge regression, models the missing values in each feature as a function of all other features and therefore, provides a more sophisticated approach than the traditional imputation methods, such as using mean, median or mode imputation.[20] After imputation, each feature was scaled to its interquartile range (IQR). IQR scaling is known to be robust to outliers and often gives better results than z-score or minmax scaling.[21] The data was then split into folds using LOHO-cv, where each iteration consists of a training fold with nine hospitals and a test fold with one hospital. The data-driven feature selection of set (6) was performed on the training fold by selecting the ten features showing the highest ANOVA F-value. Because for each iterations, the training fold consists of nine different hospitals, the selected features with the highest F-values can differ due to heterogeneity between hospitals. To be able to describe the ten most predictive features in further analysis, the features selected most often over all iterations are presented. If two feature sets are selected equally often, the set with the highest summed F-values was chosen. Both missing value imputation and feature selection were performed independently on the training and test set. After feature selection, both models were fitted and parameters optimized by a 50-iteration randomized grid search using a stratified shuffle split cross-validation. A schematic overview of all the processing steps is shown in figure 1 and the grid search parameters are shown in supplementary table 2. All code in the pipeline was implemented using the Scikit-learn python package.[18] To adhere to guidelines on transparent reporting of multivariable prediction models, the TRIPOD checklist is included in the supplementary table 3.[22] All code used in this paper is available at DOI:10.5281/zenodo.4077342

**Figure 1.**
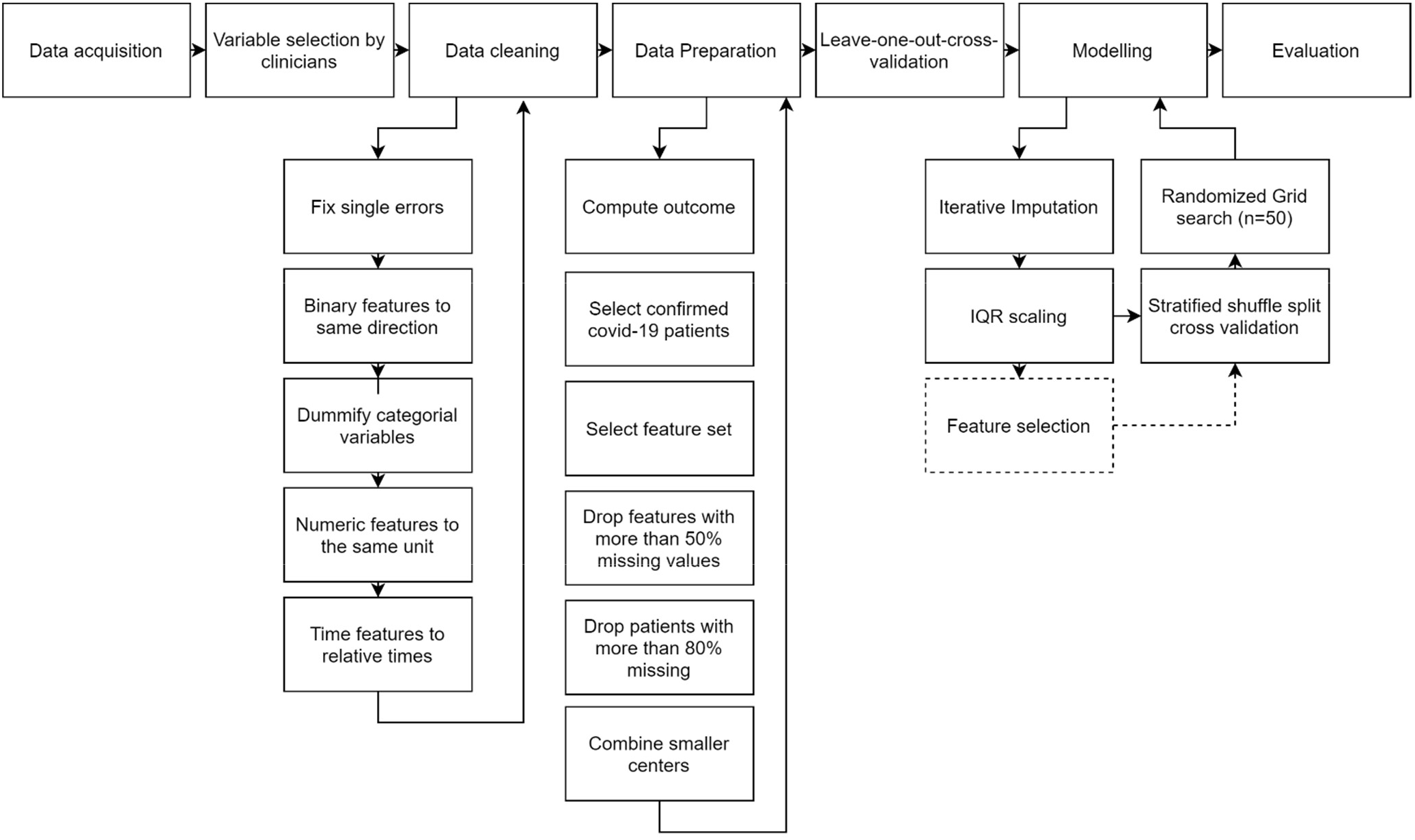
A schematic overview of all steps involved data acquisition to model evaluation. The dotted line depict the step only used during feature selection of the 10 best features.

### Performance analysis

Model performance was assessed using area under the curve (AUC), sensitivity, specificity, positive predictive value (PPV) and negative predictive value (NPV). Except for AUC, the metrics require a binary classification instead of likelihood and therefore the cutoff threshold was tuned to the shortest distance to the upper-left corner in the receiver operating curve (ROC) plot, which was named as the ‘optimal’ threshold in further analysis. In addition, a confusion matrix was derived over the complete dataset and for each center, also tuned to the optimal threshold

### Feature importance

Feature importance of models are described using SHAP (SHapley Additive exPlanations),[23] a game theoretic approach to explain the output of any machine learning model. SHAP computes the average contribution of all features by permuting all of them and subsequently evaluating the error in the prediction for when a given feature is either included or not in the model. With SHAP, the impact of low and high values of a given feature on the models’ predictions can be evaluated, as well as how impactful the feature is in predicting the correct class.[23]

### Sensitivity analysis for ICU admitted patients

During a large influx of patients suffering from life threatening lung infections, it is most likely that the ICU is to be exhausted first due to the low bed count and invasive ventilation capacity. It is therefore important to analyze whether the model also performs well on ICU admitted patients, as triage might be dependent on ICU capacity. In the Netherlands, triage was prevented by distributing patients to districts with fewer admissions or German hospitals. However, possible bias may already be present in the selection of patients, because, for example, certain patients might not be admitted to the ICU because of old age, premorbid characteristics, presentation with multi-organ failure and patients’ own treatment restraints wishes. For these reasons, both LR and XGB performances were assessed by training on the complete dataset and on ICU patient subgroup.

### Age as feature

To compare the models to clinical practice, the performance was compared with two age-based decision rules that have been applied in practice during crisis.[2] The rules were translated as follows: 1) If age is above 70 then the outcome is considered unfavorable and 2) If age is above 80 then the outcome is considered unfavorable.

Furthermore, it was assessed whether age is important for the final prediction to be able to contribute to the ongoing socio-ethical debate in the Netherlands. In July 2020, a discussion between ethicists, medical professionals and policy makers was started about criteria for triage to decide which patients receive ICU care during acute hospital care shortage. The main point of discussion was that the Dutch government was firmly opposed to using an age-based decision rule because it is in violation of the constitution, which states that everyone should be treated equal and discrimination on any ground is illegal. To contribute to this discussion, the effect of age on the best performing model was assessed, by retraining the model on the same feature set, while excluding age as feature.

## RESULTS

### Patient population

The database included 2527 patients from ten different hospitals at June 8, 2020. 223 patients were excluded because they had no recorded outcome at the time of analysis (e.g. patient was still in the hospital, but shorter than 21 days). Subsequently, 31 patients were excluded because they did not have a confirmed COVID-19 infection, resulting in 2273 patients included for modelling. Of these patients, 1195 were discharged home and not re-admitted, 76 were discharged to a nursing home and 232 were discharged to a rehabilitation unit. Furthermore, 509 patients died and 7 patients were discharged to palliative care. Of the remaining 254 patients that were still in the hospital at 21 days after admission, 112 patients were at the ward or medium care and 142 in the ICU. In total, the data included 516 unfavorable outcomes and 1757 favorable outcomes. To better balance the samples per hospital, the two smallest hospitals (n=59 and n=70) were combined. The resulting ratio of unfavorable outcome / total patients per hospital is 19% (n=261), 14% (n=169), 10% (n=118), 31% (n=317), 14% (n=113), 21% (n=401), 27% (n=325), 27% (n=440) and 19% (n=129).

### Feature description

Two features, history of smoking and alcohol abuse, were removed because of multi-interpretable questions in the rCCRF. One feature was removed in the Clinical presentation feature set and 11 features in Laboratory & Radiology findings feature set for missing more than 50% values. No patient records were excluded for missing more than 80% values. After preprocessing, Premorbid and Clinical presentation features had 2.8% and 4.0% missing values, respectively. The admission Laboratory and Radiology features showed 21.6% missing values. See supplementary table 1 and 4 for a complete overview of features and missing values. Descriptive statistics of a selection of features are shown in table 1.

**Table 1.**
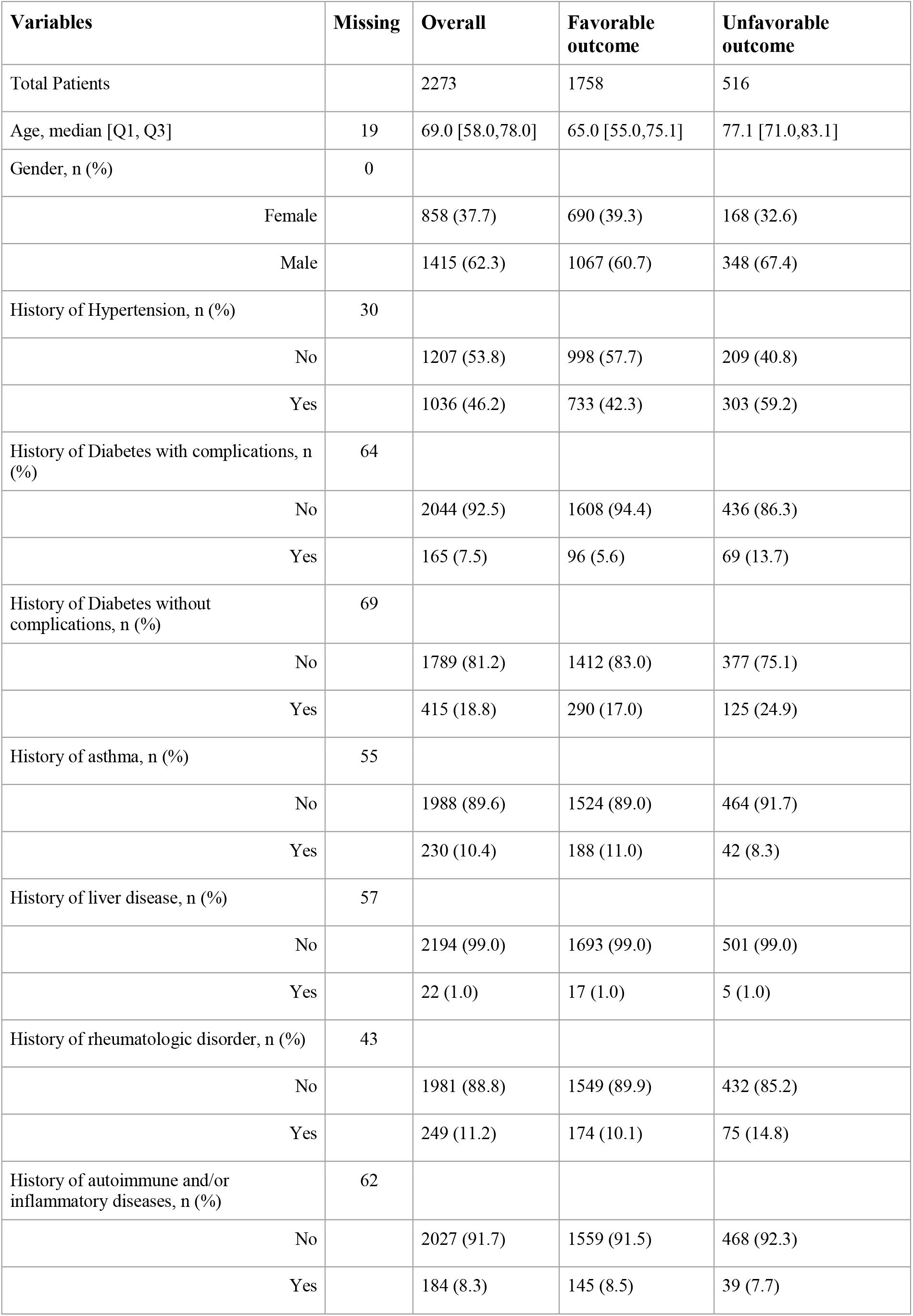
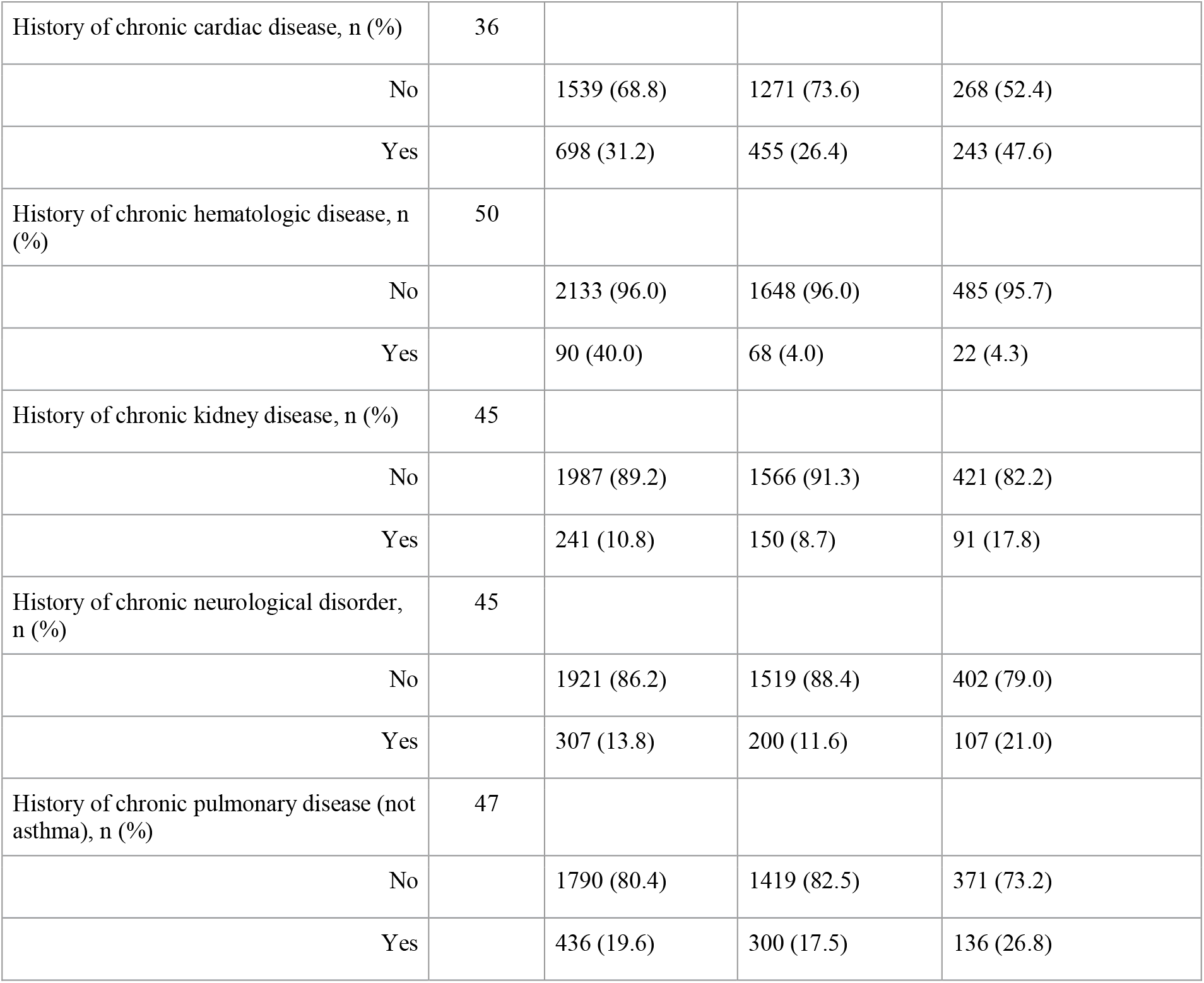
Patients characteristics per outcome group and a selection of features.

### Overall model performance

XGB and LR performed equal on the premorbid set with an AUC of 0.77 (95%-CI, 0.73 to 0.81) and 0.77 (0.72 to 0.81), respectively. On all other feature sets, XGB performed better than LR, although most 95% confidence intervals overlapped. Both XGB and LR achieved the highest AUC on the 10 best features (0.82, 0.79 to 0.85 and 0.81, 0.77 to 0.85, respectively). Figure 2A shows a comparison of the AUCs per feature set.

**Figure 2.**
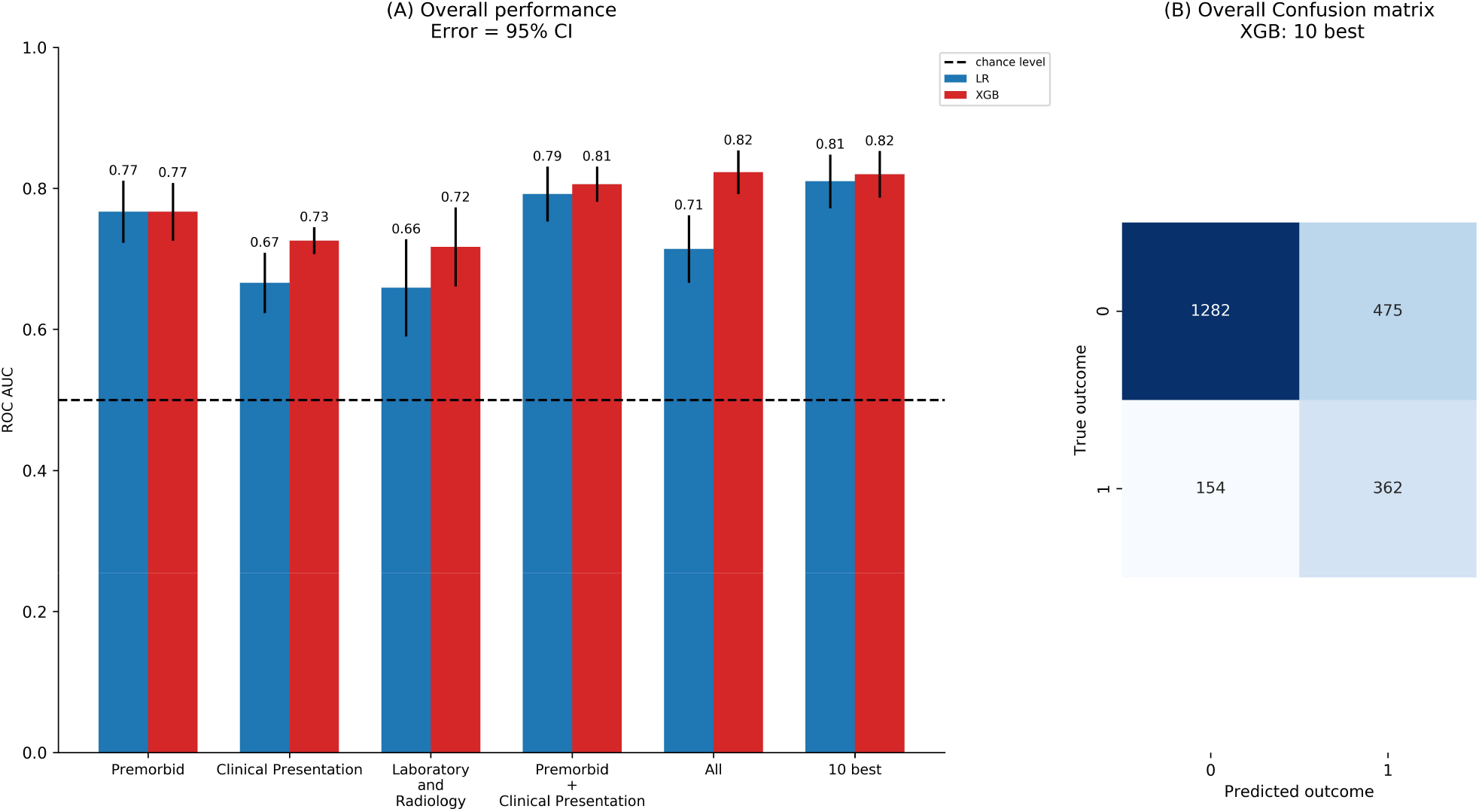
<u>Panel A</u>: Overall performance of both models per feature set. All models perform well above chance level. XGB generally performs better than LR, except on the premorbid feature set, where both models performed equal. The highest performance was achieved by XGB on both all features and the 10 selected features. <u>Panel B</u>: The confusion matrix of the best performing models, XGB trained on the 10 selected features. The prediction threshold was tuned to the shortest distance to the upper left corner of the AUC plot to create the ‘optimal’ binary prediction.

**Figure 3.**
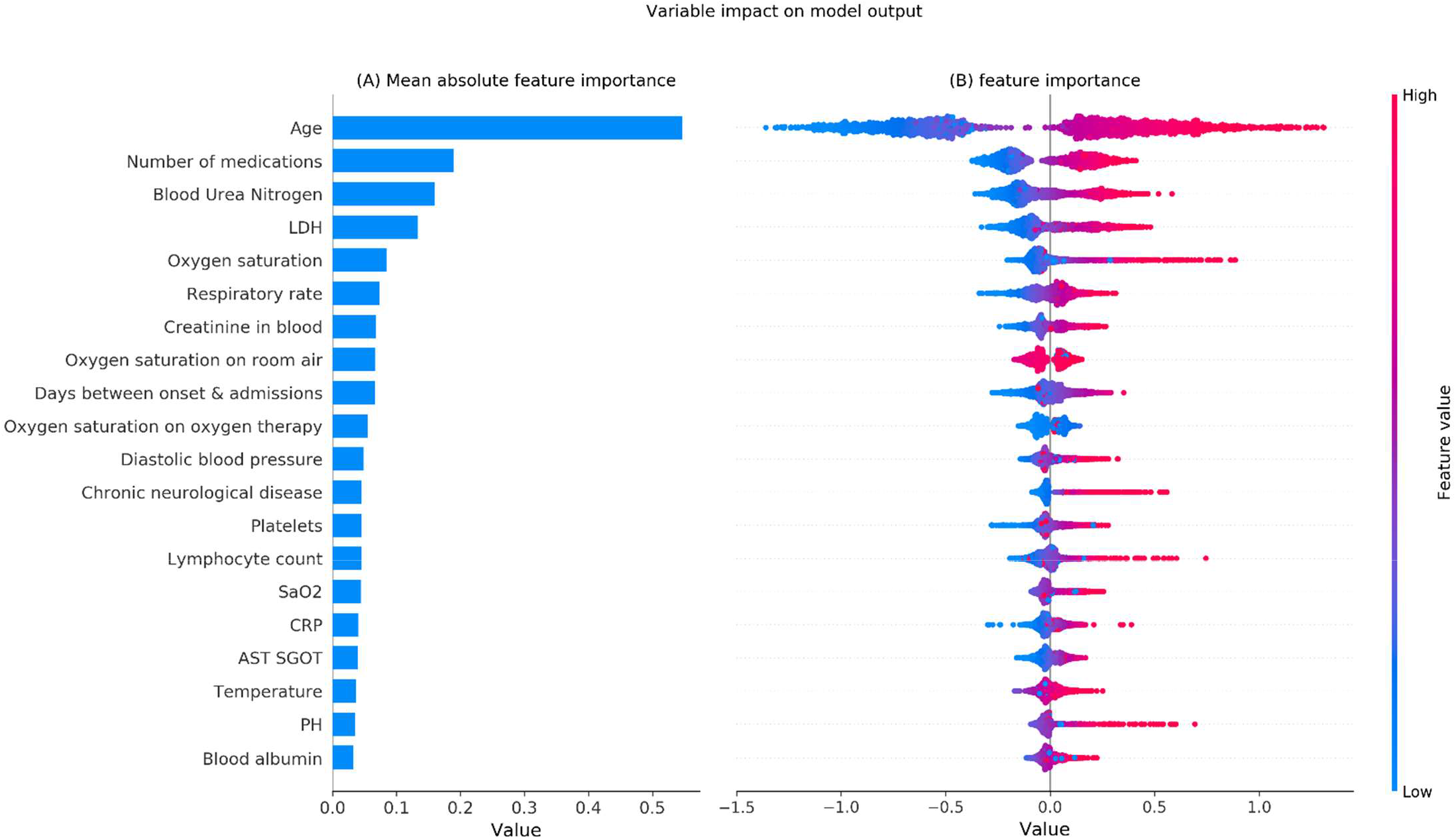
SHAP values of XGB trained on all features. To prevent readability issues, only the top 20 features are shown and the SHAP value range is set from −1.5 to 1.5, visually cutting of a few outliers. The color of each data points depicts the height of the value, where red corresponds to high values and blue to low values. SHAP values above 0 suggest a positive association with the outcome. Given the outcome is defined as mortality within 21 days, the positive SHAP values translate to association with higher mortality.

Sensitivity and specificity were comparable between the algorithms. Overall, the NPV was high and the PPV was low, as the number of patients with a favorable outcome was considerably higher than the number of patients with an unfavorable outcome. This implies that the model can make accurate predictions of favorable outcomes, but less accurate predictions of unfavorable outcomes. All results are shown in table 2. The results from internal cross-validation were comparable and shown in supplementary table 5.

**Table 2.** Evaluation metrics for both classifiers for each feature set. The average and 95% confidence intervals over all LOHO-cv iterations are presented. Values in bold represent the best performance for each metric per classifier. AUC: area under the curve; PPV: positive predictive value; NPV: negative predictive value; LR: logistic regression; XGB: extreme gradient boosting.

| Classifiers | Featureset | AUC | Sensitivity | Specificity | PPV | NPV |
| --- | --- | --- | --- | --- | --- | --- |
| LR | Premorbid | 0.77 (0.72-0.81) | 0.73 (0.61-0.84) | 0.71 (0.64-0.78) | 0.39 (0.35-0.44) | 0.91 (0.88-0.95) |
|  | Clinical Presentation | 0.67 (0.62-0.71) | 0.60 (0.51-0.68) | 0.63 (0.57-0.69) | 0.30 (0.22-0.38) | 0.86 (0.83-0.90) |
|  | Laboratory and Radiology | 0.66 (0.59-0.73) | 0.65 (0.47-0.83) | 0.54 (0.34-0.73) | 0.25 (0.16-0.34) | 0.83 (0.74-0.91) |
|  | Premorbid + Clinical Presentation | 0.79 (0.75-0.83) | 0.71 (0.62-0.80) | **0.71 (0.66-0.75)** | 0.38 (0.32-0.43) | 0.91 (0.89-0.93) |
|  | All | 0.71 (0.67-0.76) | 0.62 (0.52-0.73) | 0.70 (0.62-0.78) | 0.36 (0.28-0.44) | 0.88 (0.85-0.92) |
|  | 10 best | **0.81 (0.77-0.85)** | **0.77 (0.68-0.85)** | 0.71 (0.65-0.77) | **0.41 (0.36-0.45)** | **0.93 (0.90-0.95)** |
| XGB | Premorbid | 0.77 (0.73-0.81) | 0.68 (0.54-0.81) | 0.60 (0.39-0.82) | 0.36 (0.29-0.43) | 0.68 (0.44-0.92) |
|  | Clinical Presentation | 0.73 (0.71-0.74) | 0.69 (0.61-0.77) | 0.64 (0.59-0.69) | 0.33 (0.26-0.40) | 0.89 (0.87-0.92) |
|  | Laboratory and Radiology | 0.72 (0.66-0.77) | 0.68 (0.60-0.75) | 0.63 (0.57-0.68) | 0.31 (0.27-0.35) | 0.88 (0.84-0.92) |
|  | Premorbid + Clinical Presentation | 0.81 (0.78-0.83) | **0.76 (0.67-0.85)** | 0.62 (0.47-0.78) | 0.36 (0.29-0.44) | 0.81 (0.62-1.00) |
|  | All | **0.82 (0.79-0.85)** | 0.66 (0.54-0.78) | 0.77 (0.65-0.89) | **0.47 (0.42-0.52)** | 0.91 (0.88-0.95) |
|  | 10 best | **0.82 (0.79-0.85)** | 0.67 (0.57-0.77) | **0.75 (0.63-0.86)** | 0.44 (0.40-0.48) | **0.91 (0.88-0.94)** |

The between-hospital performance variation was small for both algorithms, shown by the small 95% confidence intervals in AUC of 0.02 to 0.06 and a low standard deviation (0.01). LR showed larger confidence intervals (0.04 to 0.07) with equal standard deviation (0.01). The small confidence intervals indicate that the models fitted the data robustly, where XGB is more robust than LR. The robustness is supported by the relatively equal ratios between correct and incorrect predictions, as shown in figure 4, which shows the confusion matrix per hospital for XGB-10 best predicting features using the optimal threshold derived from the complete dataset.

**Figure 4.**
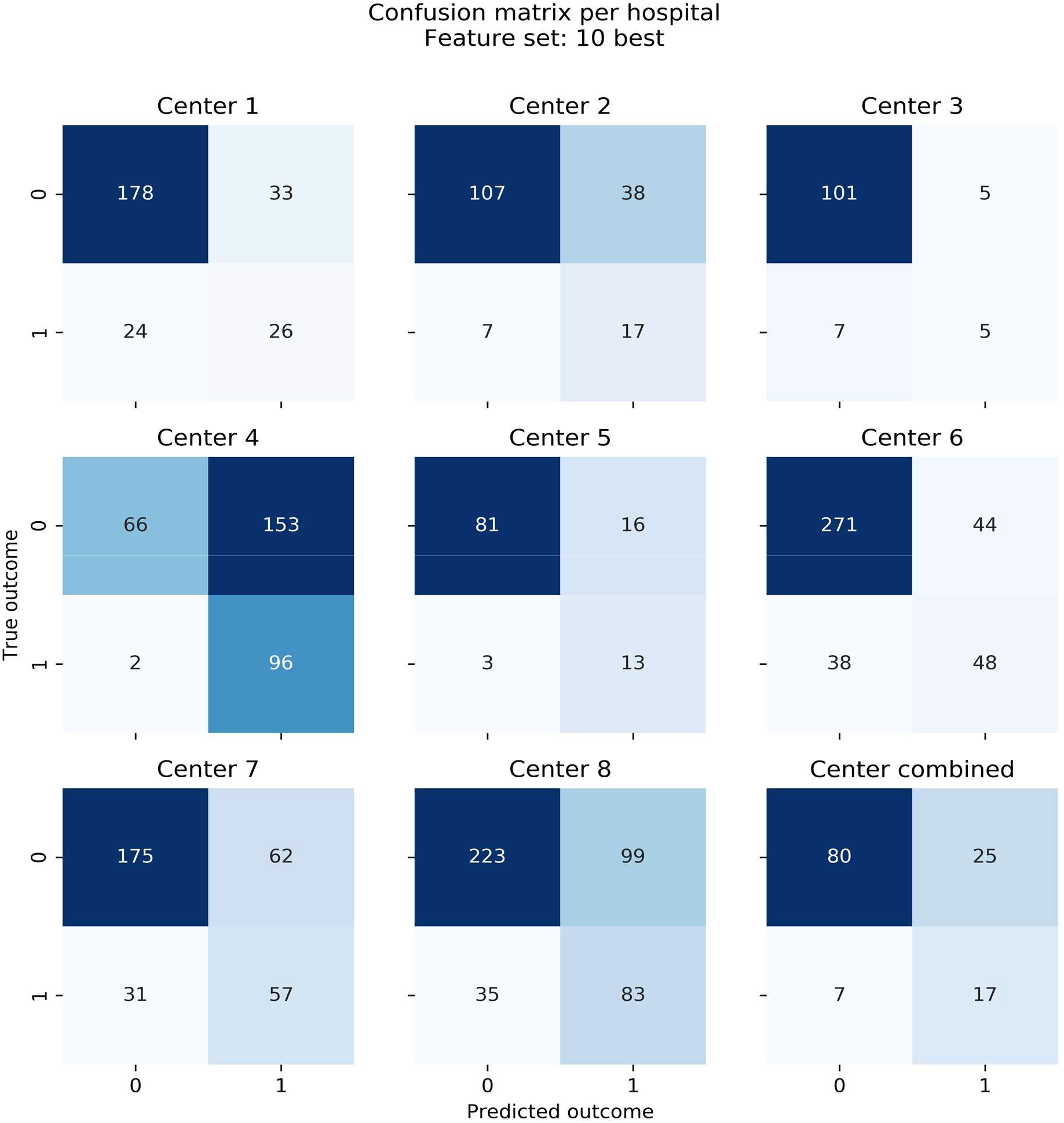
Confusion matrix per center as predicted by XGB trained on the 10 selected features. Prediction threshold is optimized by the shortest distance to upper-left corner in ROC plot of the complete dataset. All matrices show comparable distributions, though center 4 shows relatively many false positives.

### Performance stability over time

With increased duration of stay within the hospital, the uncertainty of the patients’ outcome may also increase. The patients’ chance of survival might change, because patients that have a longer hospital stay are likely to have a more complicated clinical course and/or get different types of treatments. Additionally, prolonged hospital stay simply allows more events to happen. To assess whether the models’ performance changes based on the duration of hospital stay, the patients were split per duration of stay and subsequently the performance per group was assessed. The result, presented in figure 5, shows that model performance does not deteriorate as the hospital duration increases, as the relative correct predictions remain between 0.6 and 0.9 and no trend is shown.

**Figure 5.**
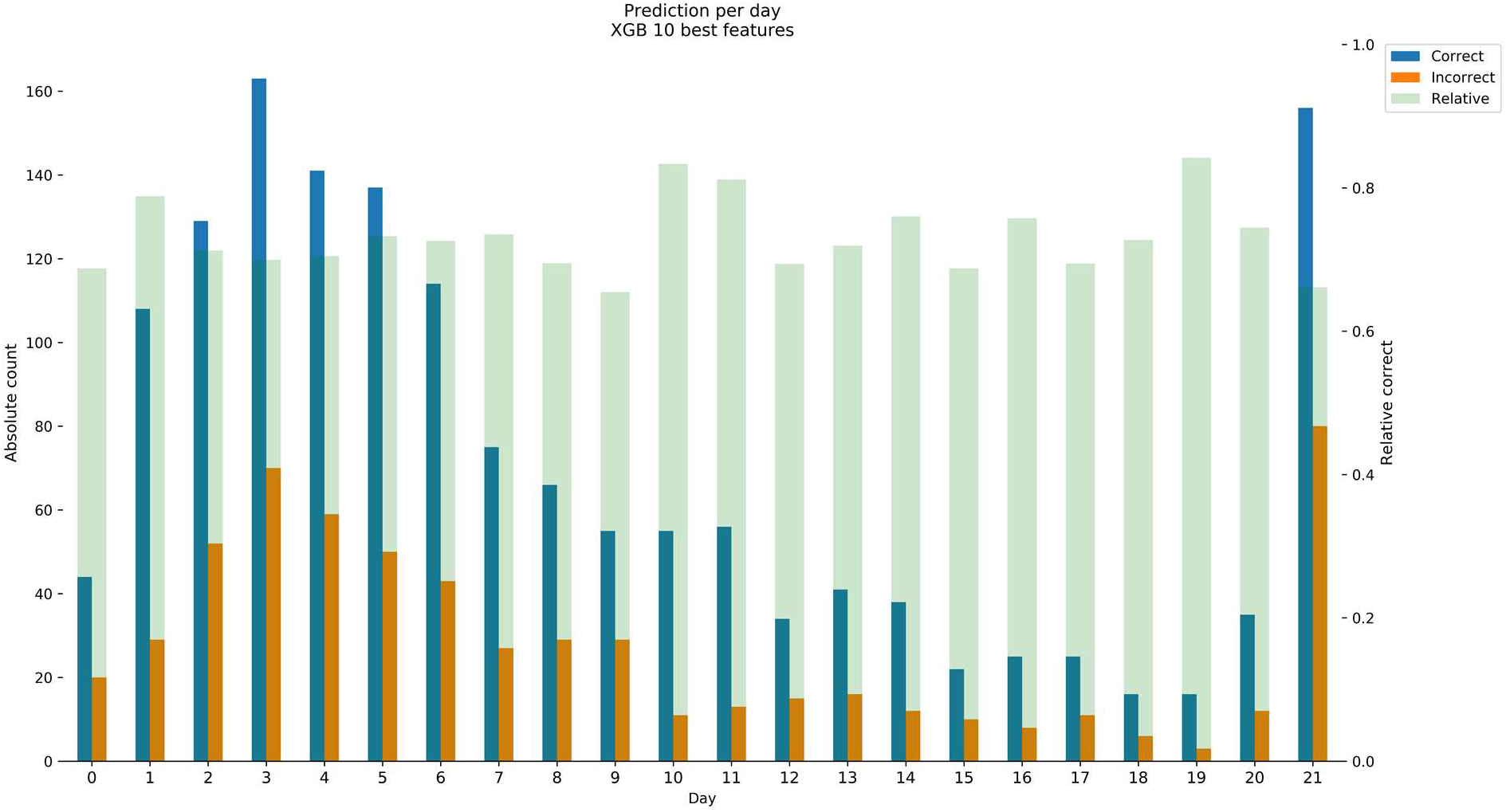
Performance per day for the XGB trained on the 10 selected features. The left y-axis shows the absolute number of correct predictions and the right y-axis the relative number correct predictions. Relative performance was calculated by *correct / (correct + incorrect)* and was well above chance level (0.5) for all days. The results indicate robust performance as the relative performance showed no decrease over time, while varying between 0.6 and 0.9. The absolute performance shows that most patients have an outcome (both favorable or unfavorable within one week after admission. A high number of patients is seen at day 21, which is caused by the aggregation of all patients that are in the hospital 21 days or longer. LR on the 10 best features shows similar performance (Figure not shown).

### Feature importance

The 10 features selected most often were, in order of highest f-value to lowest f-value: age, urea nitrogen, number of home medications, oxygen saturation (%), history of chronic cardiac disease, oxygen saturation is measured on room air, oxygen saturation is measured on oxygen therapy, blood Lactate Dehydrogenase (LDH) and blood gas pH value. The two ‘oxygen is measured on’ features are binary features that determine whether the oxygen saturation (%) is measured on room air or during oxygen therapy. The features were chosen independent of the choice of model; therefore, the selected features were the same for both LR and XGB. Figure 3A and 3B show the SHAP values per feature based on all features, for readability, only the top 20 features are shown. The features selected by the ANOVA in pre-training are also present in the top features computed be the SHAP values in post-training, which strengthens the likelihood of these features being the most important features within this dataset. This is also shown by the fact that LR scored notably higher by using the 10-best features than using all features and XGB showing equal performance using 10-best or all features.

### Sensitivity analysis for ICU patients

Of the 2273 included patients, 384 (17%) were admitted to the ICU at any time during the hospitalization. LR showed highest overall performance on ICU patients with an AUC of 0.71 (0.65 to 0.78). XGB showed the highest performance on both Premorbid and Premorbid + Clinical presentation features (0.69, 0.59 - 0.79). See table 3 for all results. For non-ICU patients, LR showed highest performance on the 10-best features (AUC 0.85, 0.81 - 0.88) and XGB on all features (AUC 0.86, 0.82 - 0.89). Compared with the results on the complete dataset, the performance dropped notably on ICU patients, decreasing in AUC by 0.04 to 0.20. The confidence intervals also increased, overall ranging from 0.03 to 0.18. Despite the discriminative power of the models decreasing, it is considered an acceptable decrease, as the initially best performing feature sets decrease only slightly and retained small confidence intervals. The decrease was expected, given that performance on a smaller subgroup is inevitably lower. In addition, the prognosis of the outcome of ICU admitted patients might change, for example, due to receiving distinct interventions only available at the ICU. When applied with caution, the models performance on ICU patients should not impede possible application in practice.

**Table 3.** Model performance on (non-)ICU subgroup. Values in bold represent the best performance per classifier per subgroup. AUC: Area under the curve, ICU: Intensive care unit, LR: Logistic regression, XGB = Extreme Gradient Boosting

| Classifiers | Feature set | AUC - ICU patients | AUC - non-ICU patients |
| --- | --- | --- | --- |
| LR | Premorbid | **0.71 (0.66-0.76)** | 0.81 (0.77-0.84) |
|  | Clinical Presentation | 0.51 (0.37-0.66) | 0.68 (0.64-0.72) |
|  | Laboratory and Radiology | 0.54 (0.45-0.63) | 0.69 (0.61-0.76) |
|  | Premorbid + Clinical Presentation | 0.60 (0.42-0.78) | 0.83 (0.80-0.86) |
|  | All | 0.63 (0.50-0.76) | 0.75 (0.72-0.79) |
|  | 10 best | 0.62 (0.44-0.80) | **0.85 (0.81-0.88)** |
| XGB | Premorbid | **0.69 (0.59-0.79)** | 0.80 (0.76-0.83) |
|  | Clinical Presentation | 0.57 (0.41-0.72) | 0.75 (0.72-0.77) |
|  | Laboratory and Radiology | 0.59 (0.52-0.66) | 0.76 (0.69-0.83) |
|  | Premorbid + Clinical Presentation | **0.69 (0.59-0.79)** | 0.84 (0.81-0.87) |
|  | All | 0.68 (0.58-0.78) | **0.86 (0.82-0.89)** |
|  | 10 best | 0.68 (0.57-0.79) | 0.85 (0.82-0.88) |

### Comparison with age-based rules for whole cohort

Of the 2273 patients, the age of 19 patients were missing and these were thus excluded for this analysis. The remaining 2254 patients, 1061 were older than 70 and 415 were older than 80. The age-based decision criteria therefore ‘predicted’ that of age > 70, 1193 will survive and 1061 will die. For age > 80 the prediction was 1839 and 415, respectively. Age > 70 showed an AUC of 0.69 (0.65 to 0.74) whereas age > 80 showed a lower AUC (0.61, 0.57 to 0.65). Figure 6 shows the confusion matrices of LR and XGB trained on the 10-best features and both age-based decision criteria. To compare both models with the age-based rules, the results were tuned to the shortest distance to the upper left corner in the ROC plot. Both LR and XGB show a higher AUC than either age-based decision criteria. The results show that the presented models can outperform earlier applied triage rules during crises and can thus provide better information based on individual medical data.

**Figure 6.**
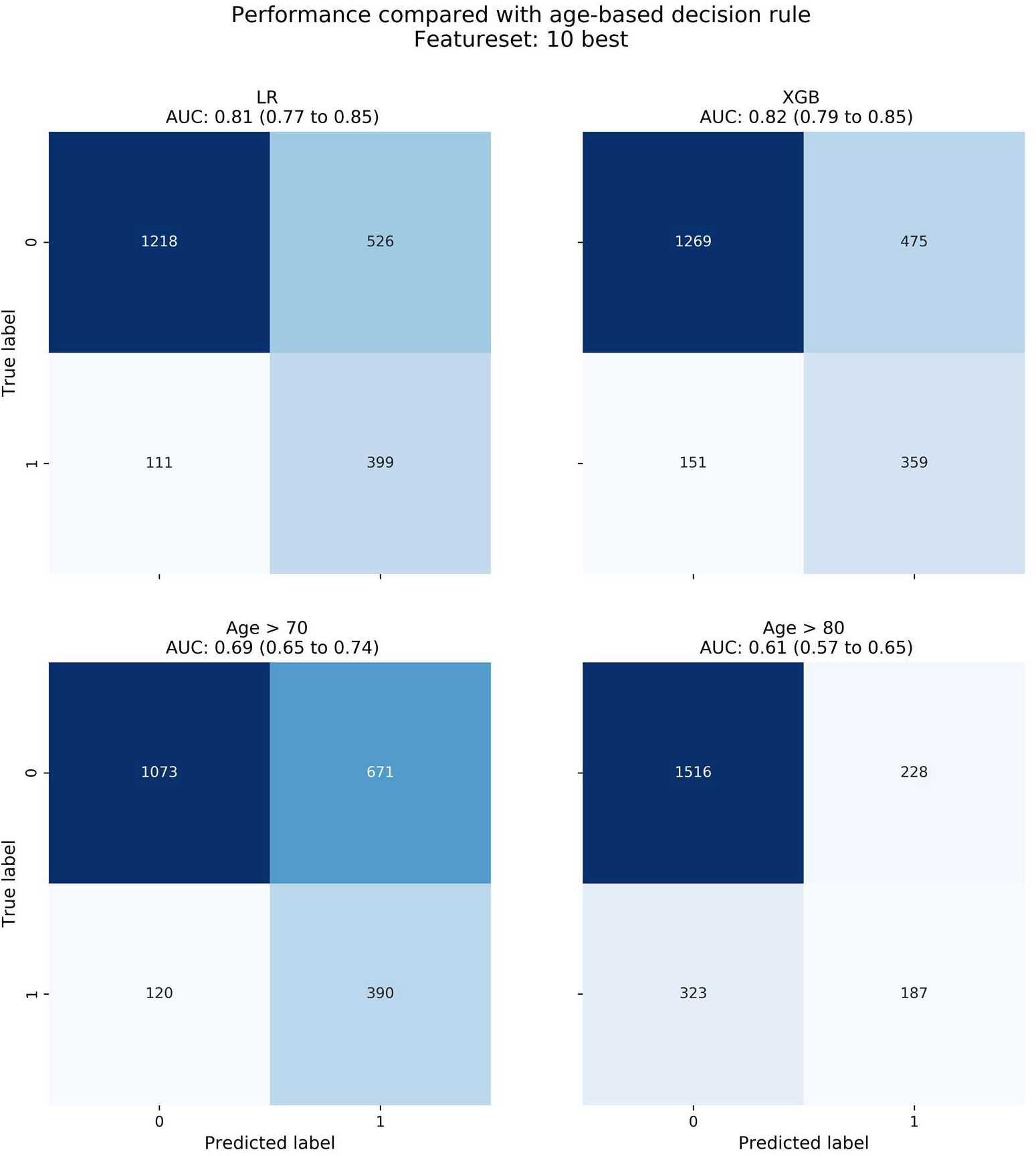
LR and XGB trained on the 10 selected features compared with two age-based decision rules. Both LR and XGB showed a higher AUC than both age-based rules. 19 patients did not have a value for age and were excluded for this analysis

### Sensitivity analysis of age as feature

The best performing model, XGB-10, was retrained an evaluated without age as feature. While expecting the performance to drop significantly, given that age was the most predictive feature by both the feature selection and SHAP analysis (Figure 3), the performance decreased only slightly from AUC of 0.82 (0.79 - 0.85) to 0.78 (0.75 - 0.81). Even though there were no signs of troublesome collinearity (supplementary figure 1), age did show high multicollinearity (variance inflation factor; VIF > 20). However, during model development, it was decided not to exclude features beforehand. Nonetheless, the high VIF indicates that the information present in age is latently present in two or more other features, which could explain the retained performance.

## DISCUSSION

The mortality of individual SARS-CoV-2 infected patients can be predicted at hospital admission with excellent performance using both linear (LR; AUC 0.81, 0.77 to 0.85) and nonlinear (XGB; 0.82, 0.79 to 0.85) models using 10 admission features that are readily available in most hospitals. Both models showed improved performance over age-based decision rules already used in practice during acute hospital bed shortage.[2] Although XGB trained on all 80 features and on the 10 best features performed comparable, the model using 10 best features may be preferred for easier translation to clinical practice.

To our best knowledge the presented models are based on one of the world’s largest multi-hospital cohorts (www.covid-predict.org) of hospitalized COVID-19 patients for which the detailed admission and clinical course has been systematically tracked by clinical personnel. The present cohort represents approximately 16% of the total hospital admissions in the Netherlands due to COVID-19 [NICE, consulted October 7th].[24] We strived to develop robust models and reduce risk of bias as much as possible, for example by adhering to recommendations by Wynants et al. [11]. Hence, there was continuous interaction between data scientists and clinicians to achieve the best of both worlds. However, our data only concerns a Dutch cohort and therefore it is uncertain if the performance remains comparable when tested on patients from other countries, where different decision-making processed could be in place. Furthermore, our models were able to make accurate predictions about favorable outcomes, given the high negative predictive value, but less accurate about unfavorable outcomes. A higher positive predictive value would reduce the amount of false positives, though this skewed performance should not hurt the main intended purpose of the model, that is when patients need to be selected based on a good estimated prognosis.

Compared to other prognostic studies, the current study improves on having not only a larger population (N=2273), but also using external validation with data from multiple hospitals. One recent study by Knight et al.[12] showed similar results using a very large multicenter cohort (n>50.000) in the United Kingdom. The authors presented two models that predicted mortality with excellent performance using comparable methodology and predictive models. Given the worldwide pressure to publish new information quickly, also leading to retracted papers in high-end journals[13][14], independent studies showing similar results is important more than ever, reducing the risk of reporting overoptimistic or spurious results

### Implications for clinicians and policymakers

Ten features were derived that were considered the most predictive features for an unfavorable outcome: age, number of home medications, admission blood values urea nitrogen/LDH/albumin, oxygen saturation (%), blood gas pH and history of chronic cardiac disease. These measures are all readily available in all Dutch and most worldwide hospitals, making the model easily applicable for prognostication or part of hospital triage tool. These factors may reflect premorbid factors (medications, albumin and cardiac history), disease severity and duration (LDH, urea nitrogen) and hypoxic and respiratory burden (oxygen saturation, pH) at hospital presentation. A social-ethical consideration is whether age should be included. Based on our results, age can be considered as evidence-based predictor in combination with other features in times of crisis and scarcity. Nevertheless, our results also indicate that the models perform only slightly worse without age as predictor, enabling model deployment when the results of a socio-ethical debate prohibit the use of age.

### Unanswered questions and future research

The current models show predictive value for hospitalized SARS-CoV-2 infected patients in a large Dutch multicenter population. The next step towards application in practice is to validate the model by testing whether the models function as expected and if they truly add value to triage. Furthermore, this paper only utilized data from the Netherlands and thus it is unknown whether the models also work on data from other countries. It might be that certain prognostic factors are country specific, related to different organizational structures or decision making in the healthcare systems.

## CONCLUSION AND RECOMMENDATION

Both LR and XGB showed excellent performance using the 10 best features, and outperformed age-based rules, with or without age included in the features. The results suggest that XGB using the 10 best features could significantly improve decision making during an acute hospital bed shortage during a COVID-19 crisis and this model is therefore recommended to be developed into a clinical tool.

## Data Availability

DATA SHARING
Not all patients provided active informed consent, and therefore data cannot be shared. The code used in this study is made publicly available and can be found at 

https://doi.org/10.5281/zenodo.4077342

## DATA SHARING

Not all patients provided active informed consent, and therefore data cannot be shared. The code used in this study is made publicly available and can be found at DOI:10.5281/zenodo.4077342

## ETHICS APPROVAL

The ethical boards of the Amsterdam University Medical Centers (20.131) and Maastricht University Medical Center approved the study protocol (MUMC: 2020-1323)

## TRANSPARENCY STATEMENT

The lead author (the manuscript’s guarantor) affirms that the manuscript is an honest, accurate, and transparent account of the study being reported; that no important aspects of the study have been omitted; and that any discrepancies from the study as originally planned (and, if relevant, registered) have been explained.

