## Supplemental material for "Predicting mortality of individual COVID-19 patients: A multicenter Dutch cohort"

**Supplementary table 1** – Individual features per feature set (adapted from the rapid COVID-19 case report form. Removed values due to missing more than 50% of values are underlined.

| **Premorbid** | **Clinical presentation** | **Laboratory / Radiology findings** |
|---|---|---|
| Age | Bleeding (Haemorrhage) | Adenovirus PCR positive |
| AIDS / HIV | Days since infection before hospital admission | ALT |
| Asthma (physician diagnosed) | Diastolic blood pressure | aPTT |
| Autoimmune and/or inflammatory diseases | Disturbed capillary refill | AST |
| Chronic cardiac disease, including congenital heart disease (not hypertension) | Heart rate | Bacteria in sputum cultured |
| Chronic hematologic disease | History of fever | Blood Albumin |
| Chronic kidney disease | Irregular heart rhythm | Blood cultures positive |
| Chronic neurological disorder | Respiratory rate | Blood Urea Nitrogen |
| Chronic pulmonary disease (not asthma) | Seizures | CO-RADS CT thorax score |
| Diabetes with complications | Shortness of breath (Dyspnea) | Blood Creatinine kinase |
| Diabetes without complications | Systolic blood pressure | Blood Creatinine value |
| Gender | Temperature | Blood CRP |
| Healthcare worker | Oxygen saturation | Blood D-dimer |
| Hypertension | Oxygen saturation measured on room air | Blood Ferritine |
| Immunosuppressive medication | Oxygen saturation on oxygen therapy | Bloor Fibrinogen |
| Malignant neoplasm | | FiO2 supplied |
| Microbiology worker | | Blood Glucose |
| Mild liver disease | | Blood Haemoglobin |
| Moderate or severe liver disease | | Influenza PCR positive |
| Number of different medicine patient uses | | Blood INR |
| Regular medicine use at home | | Blood Lactate value |
| Rheumatologic disorder | | Blood LDH |
| Rheumatologic disorder | | Other Infectious Respiratory diagnosis confirmed |
| | | PaO2 – Arterial blood gas |
| | | PaO2 – Capillary blood gas |
| | | PaO2 – Venous blood gas |
| | | PaCO2 – blood gas |
| | | PH value – blood gas |

| | | |
|---|---|---|
| | | Blood Platelets value |
| | | Blood Potassium |
| | | Blood PT |
| | | SaO2 – Blood gas |
| | | Blood Lymphocyte count |
| | | Blood Neutrophil Count |
| | | Blood Sodium |
| | | Thoracic CT findings |
| | | Blood Total Bilirubin |
| | | Blood Total calcium |
| | | Blood White blood cell count value |
| | | Presence of infiltrates on lung imaging |

**Supplementary table 2:** Hyper-parameters used for optimizing LR or XGB models. A 50-iteration randomized grid-search was used for both models. The values in square brackets under parameters name are the parameter names as per function in the code.

| Classifier | Parameter Name | Parameter Value |
|---|---|---|
| **XGB** | [Learning rate] | [0.1, 0.01, 0.001] |
| | [Gamma] Minimum loss reduction required to make a further partition on a leaf node of the tree | [0.1, 0.01, 0.001] |
| | [N estimators] | [100, 200, 300, 500, 700] |
| | [Subsample] Subsample ratio of the training instances | [0.5, 0.7, 0.9] |
| | [Colsample by tree] Parameters for subsampling the columns | [0.5, 0.6, 0.7, 0.8] |
| | [Max depth] Maximum depth of a tree | [2, 4, 6, 8] |
| **LR** | [Solver] | [Saga] |
| | [Penalty] Regularization penalty type | [Elasticnet, L2] |
| | [L1 Ratio] Ratio between L1 and L2 regularization | [0.0, 0.2, 0.4, 0.6, 0.8, 1.0] |
| | [C] Inverse regularization strength | [0.0, 0.001, 0.006, 0.046, 0.359, 2.783, 21.544, 166.81, 1291.55, 10000.0] |

**Supplementary table 3** – Tripod checklist, the page columns states the page where the information can be found. appx: appendix, suppl: supplementary

| Section/Topic | Item | Checklist Item | Page |
|---|---|---|---|
| **Title and abstract** | | | |
| **Title** | 1 | Identify the study as developing and/or validating a multivariable prediction model, the target population, and the outcome to be predicted. | 1 |
| **Abstract** | 2 | Provide a summary of objectives, study design, setting, participants, sample size, predictors, outcome, statistical analysis, results, and conclusions. | 2 |
| **Introduction** | | | |
| **Background and objectives** | 3a | Explain the medical context (including whether diagnostic or prognostic) and rationale for developing or validating the multivariable prediction model, including references to existing models. | 3 |
| | 3b | Specify the objectives, including whether the study describes the development or validation of the model or both. | 3, 4 |
| **Methods** | | | |
| **Source of data** | 4a | Describe the study design or source of data (e.g., randomized trial, cohort, or registry data), separately for the development and validation data sets, if applicable. | 5 |
| | 4b | Specify the key study dates, including start of accrual; end of accrual; and, if applicable, end of follow-up. | 5 |
| **Participants** | 5a | Specify key elements of the study setting (e.g., primary care, secondary care, general population) including number and location of centres. | 5 |
| | 5b | Describe eligibility criteria for participants. | 5 |
| | 5c | Give details of treatments received, if relevant. | n/a |
| **Outcome** | 6a | Clearly define the outcome that is predicted by the prediction model, including how and when assessed. | 5, 6 |
| | 6b | Report any actions to blind assessment of the outcome to be predicted. | n/a |
| **Predictors** | 7a | Clearly define all predictors used in developing or validating the multivariable prediction model, including how and when they were measured. | 6, table 2, suppl table 1, suppl table 4 |
| | 7b | Report any actions to blind assessment of predictors for the outcome and other predictors. | n/a |
| **Sample size** | 8 | Explain how the study size was arrived at. | 5 |

| | | | |
|---|---|---|---|
| **Missing data** | 9 | Describe how missing data were handled (e.g., complete-case analysis, single imputation, multiple imputation) with details of any imputation method. | 7 |
| **Statistical analysis methods** | 10c | For validation, describe how the predictions were calculated. | 7, figure 1 |
| | 10d | Specify all measures used to assess model performance and, if relevant, to compare multiple models. | 8 |
| | 10e | Describe any model updating (e.g., recalibration) arising from the validation, if done. | n/a |
| **Risk groups** | 11 | Provide details on how risk groups were created, if done. | n/a |
| **Development vs. validation** | 12 | For validation, identify any differences from the development data in setting, eligibility criteria, outcome, and predictors. | 7, 8 |
| **Results** | | | |
| **Participants** | 13a | Describe the flow of participants through the study, including the number of participants with and without the outcome and, if applicable, a summary of the follow-up time. A diagram may be helpful. | 9 |
| | 13b | Describe the characteristics of the participants (basic demographics, clinical features, available predictors), including the number of participants with missing data for predictors and outcome. | 10, Table 1 |
| | 13c | For validation, show a comparison with the development data of the distribution of important variables (demographics, predictors and outcome). | Table 1 |
| **Model performance** | 16 | Report performance measures (with CIs) for the prediction model. | 11, 12, 13, table 1, table 2, table 3, Figure 1, suppl table 4 |
| **Model-updating** | 17 | If done, report the results from any model updating (i.e., model specification, model performance). | n/a |
| **Discussion** | | | |
| **Limitations** | 18 | Discuss any limitations of the study (such as nonrepresentative sample, few events per predictor, missing data). | 17 |
| **Interpretation** | 19a | For validation, discuss the results with reference to performance in the development data, and any other validation data. | 17 |
| | 19b | Give an overall interpretation of the results, considering objectives, limitations, results from similar studies, and other relevant evidence. | 19 |
| **Implications** | 20 | Discuss the potential clinical use of the model and implications for future research. | 18 |
| **Other information** | | | |
| **Supplementary information** | 21 | Provide information about the availability of supplementary resources, such as study protocol, Web calculator, and data sets. | appx |

| | | | |
|---|---|---|---|
| **Funding** | 22 | Give the source of funding and the role of the funders for the present study. | 17 |

**Supplementary table 4** – Feature set description before and after preprocessing. The average absolute missing, average relative missing, minimum missing per feature and maximum missing per feature are described after preprocessing. The Laboratory & Radiology feature set shows notably higher amount of missing values, likely because no all laboratory values are measured for each patient. In addition, some features were added at a later stage in development, resulting in missing values for patients already included. Note that the difference in features before and after preprocessing includes feature removal and dummification, which occasionally increases the amount of features (see the Clinical Presentation feature set).

| **Feature set** | **N features** | **N Features after preprocessing** | **Average absolute missing** | **Average relative missing (%)** | **Min missing** | **Max missing** |
|---|---|---|---|---|---|---|
| **Premorbid** | 24 | 22 | 63 | 2.8 | 0 | 349 |
| **Clinical presentation** | 14 | 15 | 92 | 4.0 | 0 | 336 |
| **Laboratory & radiology** | 42 | 33 | 490 | 21.6 | 0 | 1023 |
| **Premorbid + Clinical presentation** | 38 | 37 | 75 | 3.3 | 0 | 349 |
| **All** | 80 | 70 | 271 | 11.9 | 0 | 1023 |

**Supplementary table 5:** Performance on internal validation. Performance is evaluated by using 10-fold random subsampling cross-validation. For each metric, the best performance per classifier is highlighted by bold text. LR performed best on all metrics except specificity when trained on the 10 selected features. XGB performed highest using all features on all metrics except negative predictive value. LR: Logistics regression, XGB: Extreme gradient boosting, AUC: Area under de curve, PPV: Positive predictive value, NPV: Negative predictive value

| Classifiers | Featureset | AUC | Sensitivity | Specificity | PPV | NPV |
|---|---|---|---|---|---|---|
| LR | Premorbid | 0.76 (0.75-0.77) | 0.75 (0.73-0.78) | 0.67 (0.65-0.70) | 0.40 (0.38-0.42) | 0.90 (0.90-0.91) |
| | Clinical Presentation | 0.68 (0.65-0.71) | 0.59 (0.50-0.68) | 0.67 (0.60-0.74) | 0.34 (0.30-0.38) | 0.86 (0.85-0.87) |
| | Laboratory and Radiology | 0.67 (0.66-0.69) | 0.59 (0.56-0.61) | 0.67 (0.64-0.71) | 0.34 (0.32-0.36) | 0.85 (0.84-0.86) |
| | Premorbid + Clinical Presentation | 0.78 (0.77-0.79) | 0.73 (0.66-0.79) | 0.72 (0.68-0.76) | 0.42 (0.39-0.46) | 0.90 (0.89-0.92) |
| | All | 0.59 (0.55-0.64) | 0.29 (0.14-0.44) | **0.83 (0.75-0.92)** | 0.35 (0.31-0.38) | 0.80 (0.77-0.83) |
| | 10 best | **0.81 (0.80-0.82)** | **0.79 (0.77-0.80)** | 0.71 (0.69-0.73) | **0.44 (0.42-0.47)** | **0.92 (0.91-0.93)** |
| XGB | Premorbid | 0.77 (0.76-0.79) | 0.78 (0.75-0.81) | 0.68 (0.66-0.70) | 0.39 (0.37-0.42) | 0.68 (0.44-0.92) |
| | Clinical Presentation | 0.73 (0.72-0.74) | 0.69 (0.67-0.72) | 0.66 (0.64-0.68) | 0.37 (0.36-0.39) | 0.89 (0.87-0.92) |
| | Laboratory and Radiology | 0.73 (0.71-0.74) | 0.70 (0.67-0.72) | 0.63 (0.60-0.66) | 0.35 (0.33-0.38) | 0.88 (0.84-0.92) |
| | Premorbid + Clinical Presentation | 0.81 (0.80-0.82) | 0.77 (0.75-0.79) | 0.71 (0.70-0.73) | 0.45 (0.43-0.47) | 0.81 (0.62-1.00) |
| | All | **0.83 (0.81-0.84)** | **0.78 (0.75-0.80)** | **0.74 (0.72-0.77)** | **0.47 (0.45-0.49)** | 0.91 (0.88-0.95) |
| | 10 best | 0.81 (0.80-0.82) | 0.75 (0.72-0.77) | 0.74 (0.71-0.76) | 0.46 (0.43-0.49) | **0.91 (0.88-0.94)** |

**Supplementary figure 1** – Pearson correlation matrix of all features. No troublesome collinearity was seen.

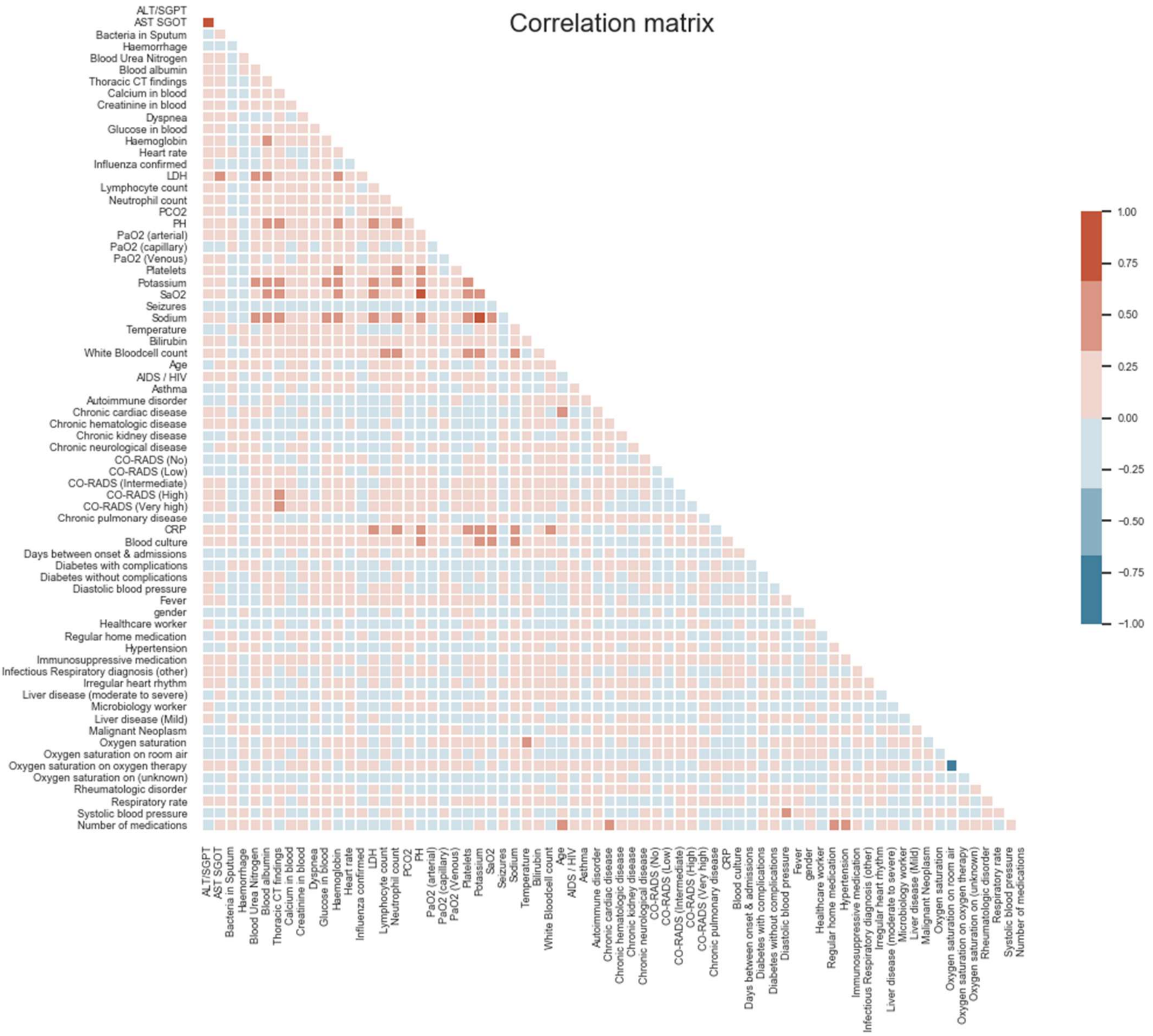
